# Multi-tissue transcriptome-wide association study identifies genetic mechanisms underlying endometrial cancer susceptibility

**DOI:** 10.1101/2020.12.03.20243758

**Authors:** Pik-Fang Kho, Gabriel Cuellar Partida, Thilo Dörk, Ellen L. Goode, Diether Lambrechts, Rodney J. Scott, Endometrial Cancer Association Consortium, Amanda B. Spurdle, Tracy A. O’Mara, Dylan M. Glubb

## Abstract

Genome-wide association studies (GWAS) of endometrial cancer have identified 16 genetic susceptibility loci. To identify candidate endometrial cancer susceptibility genes, we have performed the first transcriptome-wide association study (TWAS) of endometrial cancer. For this analysis, we have used the largest endometrial cancer GWAS and gene expression from 48 tissues to maximise statistical power. We used colocalisation analysis to prioritise seven candidate susceptibility genes, including *CYP19A1*, which wes previously established as an endometrial cancer susceptibility gene. Notably, one of the candidate susceptibility genes was located at 17q24.2, a potentially novel risk locus for endometrial cancer. Using phenome-wide association analysis, candidate susceptibility genes were found to associate with traits related to endometrial cancer risk factors, in addition to other clinical phenotypes that may provide novel risk factors. TWAS data were also used to perform drug repurposing analysis and identified 14 compounds; two of these are tubulin inhibitors, a drug class used to treat advanced endometrial cancer. In summary, this study has revealed biologically relevant endometrial cancer susceptibility genes, providing insights into endometrial cancer aetiology and avenues for the development of new treatments.

## Introduction

Endometrial cancer is the most commonly diagnosed gynaecological cancer in developed countries, with incidence expected to continue to increase over the next decade (reviewed in (Morice, Leary, Creutzberg, Abu-Rustum, & Darai, 2016)). To explore the role of common genetic variation in endometrial cancer susceptibility, we recently conducted a genome-wide association study (GWAS) meta-analysis and identified 16 susceptibility loci (O’Mara et al., 2018). Despite the success of GWAS in identifying genetic variants underlying endometrial cancer risk, biological interpretation remains challenging given that the majority of the GWAS risk variation is located in non-protein-coding regions (reviewed in (O’Mara, Glubb, Kho, Thompson, & Spurdle, 2019)).

Trait-associated loci identified by GWAS have demonstrated enrichment for expression quantitative trait loci (eQTLs), suggesting that changes in gene expression link genetic variation and phenotypes (Consortium et al., 2017). As eQTL data are not typically available for GWAS samples, the transcriptome-wide association study (TWAS) approach has been developed to integrate eQTL and GWAS data from independent sample sets (Gamazon et al., 2015). TWAS uses effect estimates from eQTLs to impute gene expression in GWAS samples, and subsequently associate the imputed gene expression with phenotypes. TWAS has an additional advantage of alleviating the multiple testing penalty of GWAS in statistical inference by testing the imputed expression of thousands of genes rather than millions of genetic variants.

TWAS has identified candidate causal genes for many complex traits, using eQTL data from disease-relevant tissues or cell types (Bien et al., 2019; Gusev et al., 2019; Lamontagne et al., 2018; Wu et al., 2018). However, endometrial or uterine eQTL data are less commonly available than for other tissues or cell types (e.g. whole blood). Nevertheless, Mortlock et al. (2020) have shown that 85% of eQTLs found in the endometrium are shared across multiple tissues, suggesting that data from other tissues could be incorporated to perform endometrial cancer TWAS. Recently, innovative multi-tissue TWAS approaches that leverage shared eQTLs across multiple tissues have become available, and provide increased statistical power to identify candidate causal genes (Barbeira et al., 2019; Hu et al., 2019). These multi-tissue TWAS approaches account for the correlation of gene expression between tissues and perform multivariate regression to predict gene expression from multiple tissues simultaneously.

In the current study, we identified candidate endometrial cancer susceptibility genes by conducting multi-tissue TWAS analysis, integrating GWAS data from the Endometrial Cancer Association Consortium (O’Mara et al., 2018) and a large-scale cis-eQTL mapping study across 48 tissues from the Genotype-Tissue Expression (GTEx) project (Consortium et al., 2017). We also performed a phenome-wide association analysis to uncover traits that relate to candidate endometrial cancer susceptibility genes and provide insights into endometrial cancer aetiology. Finally, we explored drug repurposing opportunities for endometrial cancer by analysing TWAS findings using the Connectivity Map database (Subramanian et al., 2017) and Open Target platform (Carvalho-Silva et al., 2019).

## Results

Multi-tissue TWAS analysis using S-MultiXcan revealed 20 genes whose imputed expression associated with endometrial cancer susceptibility (FDR<0.05), five of which (*BHLHE41, CYP19A1, EIF2AK4, IQSEC1* and *SRCIN1*) also passed a Bonferroni correction threshold (P<2×10^−6^) (**Table 1, Figures 1 and 2**). Given that the TWAS associations could result from linkage disequilibrium (LD) between genetic variants independently affecting gene expression and endometrial cancer risk, we performed colocalization analysis to reduce LD contamination (Giambartolomei et al., 2014). The associations of *AC145343*.*2, BHLHE41, CYP19A1, RAB11FIP4, RP11-521C20*.*2, SRCIN1* and *SKAP1* were supported by colocalization of eQTL and GWAS signals (**Table 1 and Figure 3**); thus, these seven genes were prioritised as candidate endometrial cancer susceptibility genes. Notably, *AC145343*.*2* is located further than 1 Mb from any previously identified endometrial cancer GWAS risk locus and may provide a novel risk region.

**Table 1.**
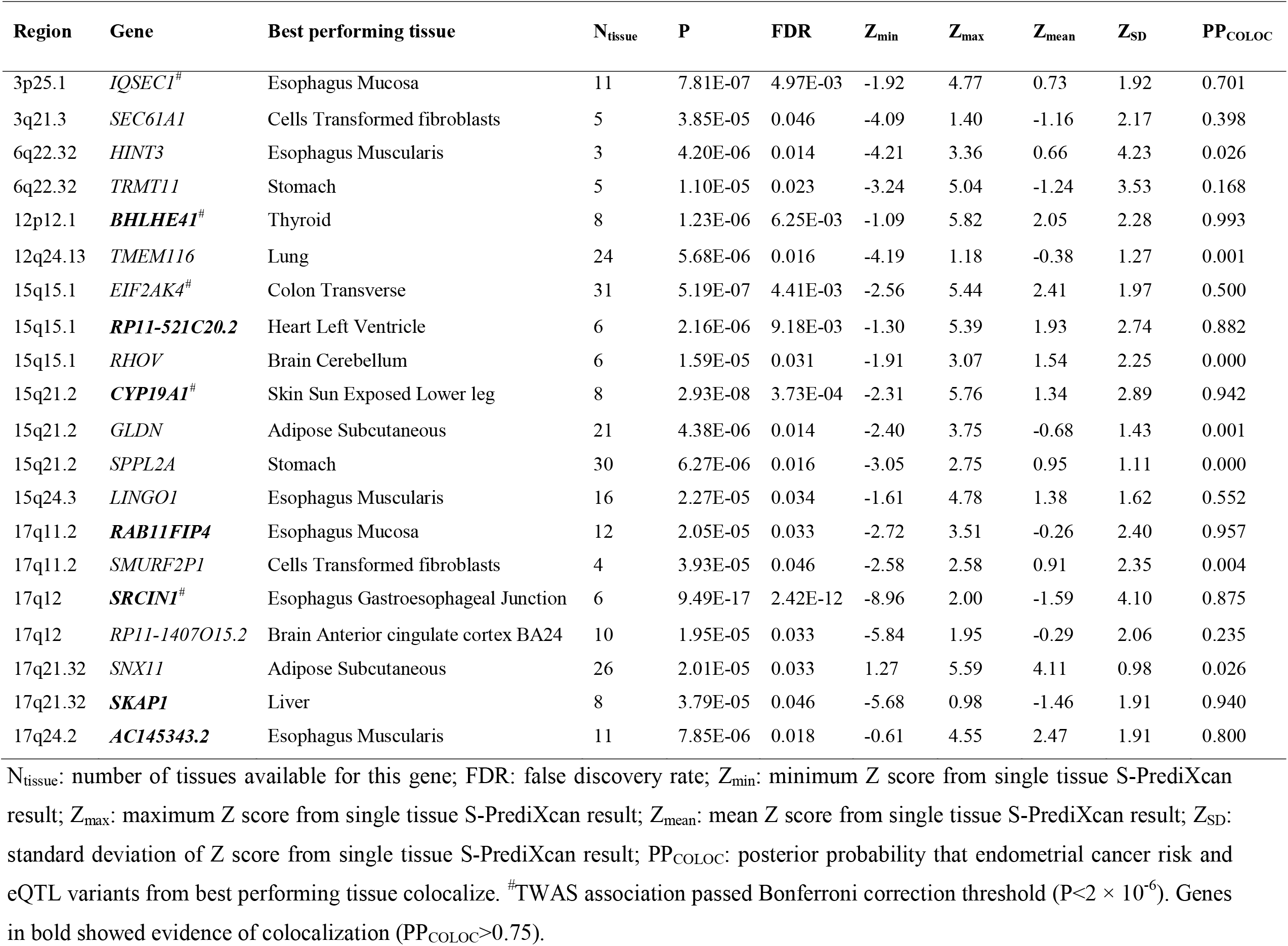
Genes associated with endometrial cancer risk as identified by TWAS (FDR<0.05)

**Figure 1.**
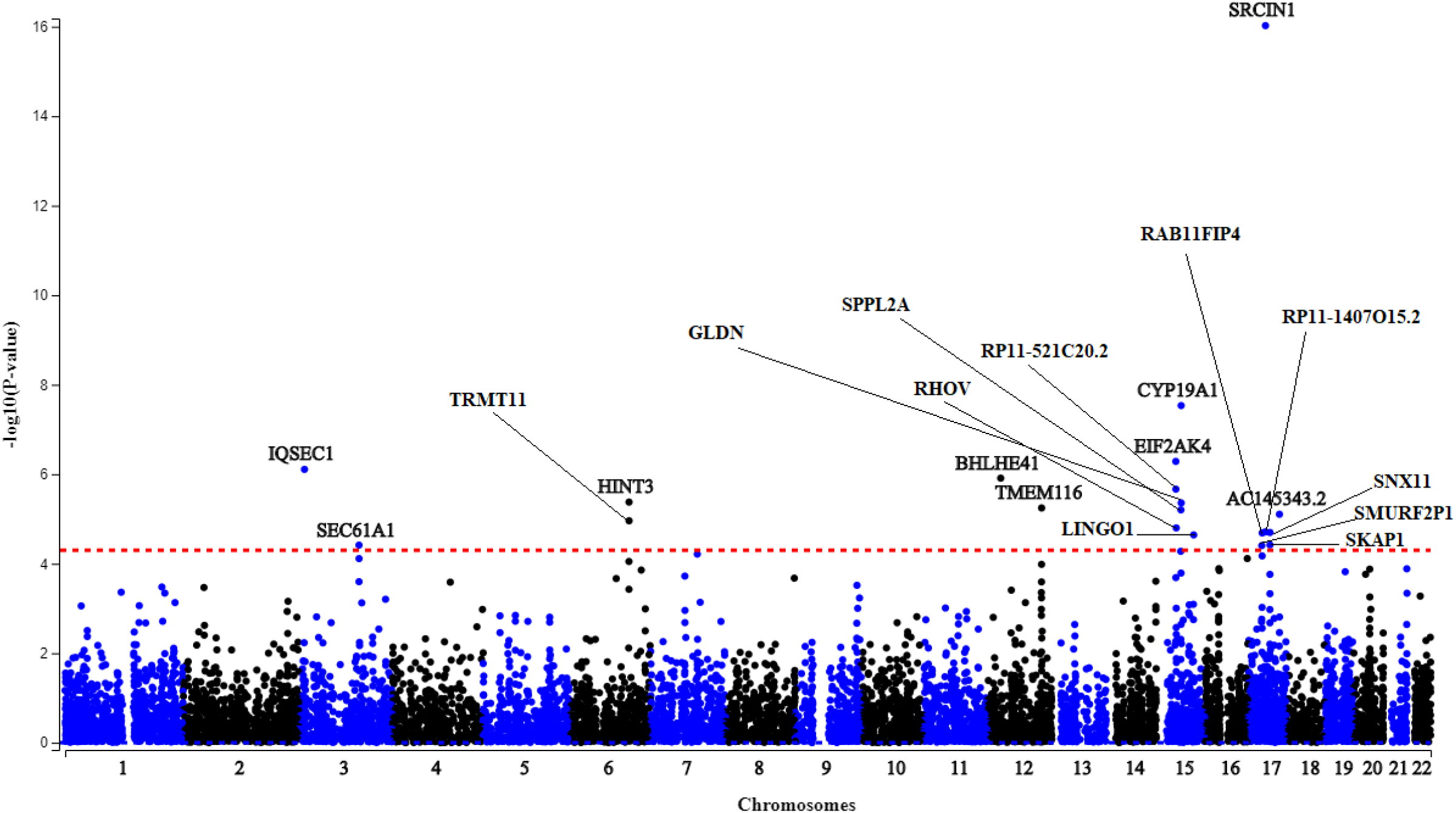
Manhattan plot of TWAS analysis of endometrial cancer risk. Genes are plotted according to chromosome and position against −log_10_(P value) of the TWAS association. Red dotted lines represent the Benjamini-Hochberg significance threshold (FDR<0.05), which is equivalent to P<5 ×10^−5^.

**Figure 2.**
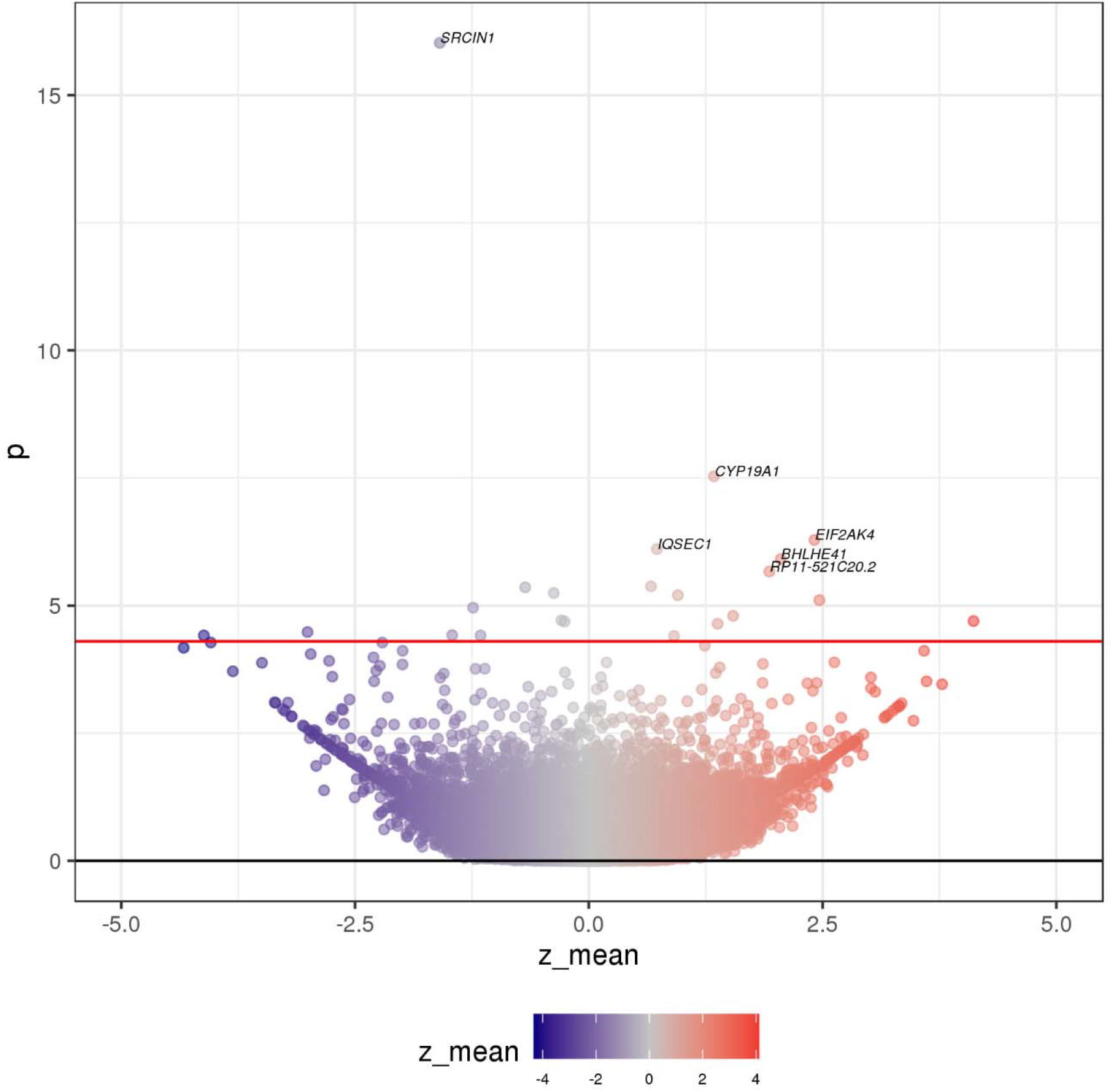
Volcano plot from TWAS analysis of endometrial cancer risk. Each dot represents the effect of the average predicted expression of each gene on endometrial cancer risk. Red line represents the Benjamini-Hochberg significance threshold (FDR< 0.05). Genes with FDR<0.01 are labelled.

**Figure 3.**
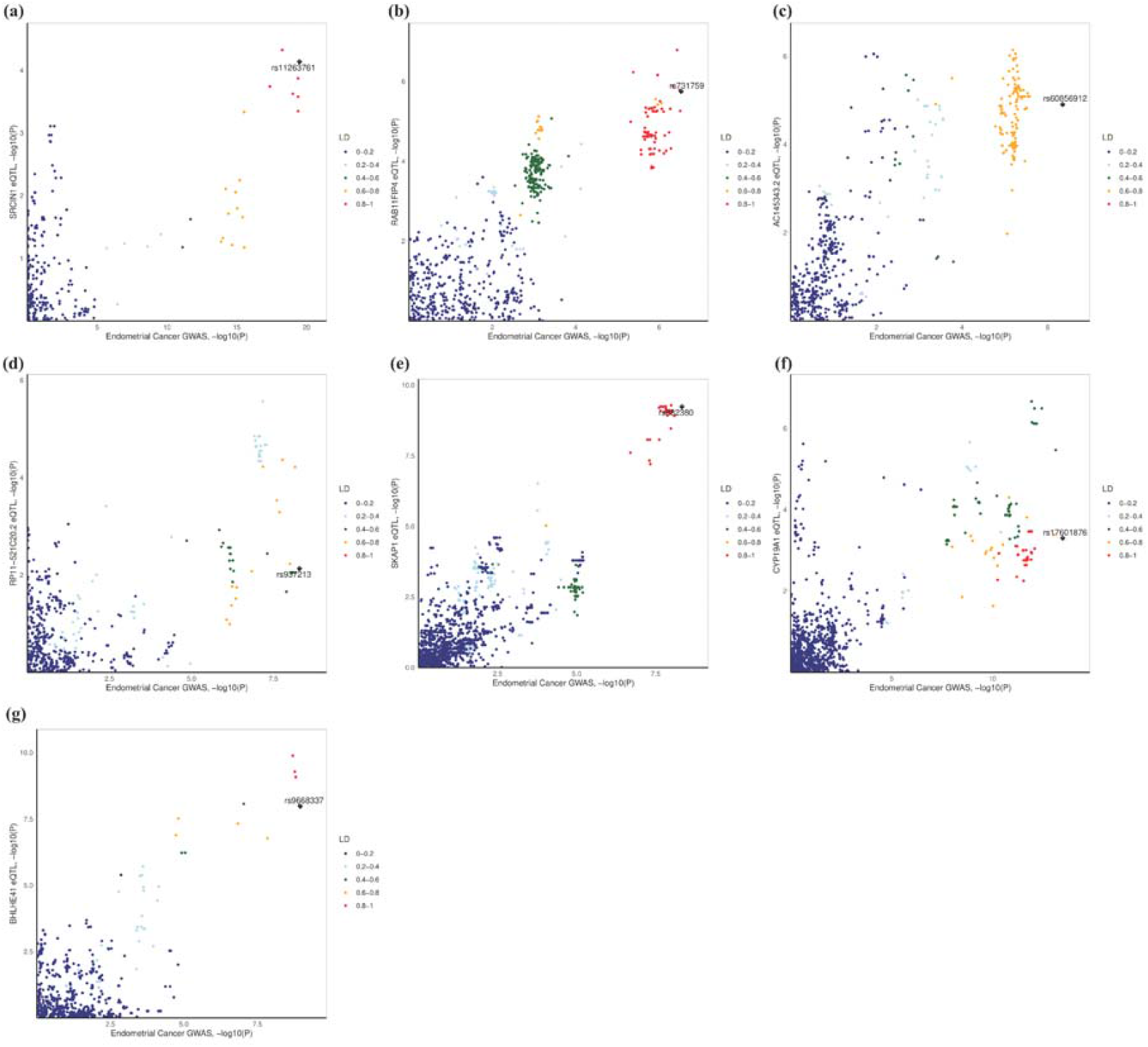
Colocalization of association signals between endometrial cancer GWAS and eQTL data from the corresponding best performing tissue in the TWAS analysis. -log10(P) of endometrial cancer GWAS was plotted against −log10(P) of eQTLs for (a) *SRCIN1*, (b) *RAB11FIP4*, (c) *AC145343*.*2*, (d) *RP11-521C20*.*2*, (e) *SKAP1*, (f) *CYP19A1* and (g) *BHLHE41*. The lead variant of each locus identified in endometrial cancer GWAS is shown as a purple diamond. Other variants are coloured according to their degree of LD with lead variant in the locus.

Secondary TWAS analysis of endometrioid endometrial cancer risk identified eight genes (FDR<0.05), seven of which were identified in the main analysis (**Supplementary Table 1 and Supplementary Figure 1**). Additionally, this analysis identified a long non-coding RNA (lncRNA), *RP11-358N4*.*6*, which was supported by eQTL colocalization but was not found in the main analysis. *RP11-358N4*.*6* is also located further than 1Mb from any previously identified endometrial cancer GWAS risk locus but was not prioritised for further follow-up as it was only identified through a male-specific tissue (i.e. testis).

Phenome-wide analysis of the seven candidate susceptibility genes highlighted three genes (*CYP19A1, AC145343*.*2* and *RAB11FIP4*) with effects on traits which were grouped into five phenotypic categories: bone, cardiovascular, haematopoiesis, diabetes, sex hormones and liver function (**Supplementary Table 2**). Three of these categories (bone, hormone and diabetes) contain traits that relate to or include genetically established endometrial cancer risk factors (e.g. hyperinsulinemia, circulating estrogen and age at menopause) (O’Mara, Glubb, et al., 2019).

We integrated data from the main TWAS analysis with drug-induced gene expression profiles from the Connectivity Map database and identified 14 compounds with profiles opposing the expression of genes associated with endometrial cancer risk (**Table 2**). These drugs included two tubulin inhibitors, ten ATPase inhibitors, a calcium channel activator and an opioid receptor antagonist. None of the 14 drugs have been approved for endometrial cancer treatment but the tubulin inhibitor vincristine has shown modest activity in a Phase II endometrial cancer study (Broun, Blessing, Eddy, & Adelson, 1993). We also explored drug repurposing opportunities for endometrial cancer using the Open Targets platform to assess the seven candidate susceptibility genes. Only the protein encoded by *CYP19A1* (aromatase), is targeted by drugs that have received clinical approval, with indications for breast cancer and Cushing syndrome (**Table 3**).

**Table 2.** Connectivity Map compounds with gene expression profiles opposing endometrial cancer risk TWAS associations.

| Connectivity score* | Drug | Drug class | Clinically tested (highest phase) or FDA approved | Indication |
| --- | --- | --- | --- | --- |
| -95.60 | nocodazole | Tubulin inhibitor | No | - |
| -95.34 | vincristine | Tubulin inhibitor | Approved | Acute lymphocytic leukemia, acute myeloid leukemia, Hodgkin and non-Hodgkin lymphoma |
| -94.75 | strophanthidin | Na<sup>+</sup>/ K<sup>+</sup> -ATPase inhibitor | No | - |
| -94.45 | cinacalcet | Calcium channel activator | Approved | Hypercalcemia and secondary hyperparathyroidism |
| -94.40 | digitoxigenin | Na<sup>+</sup>/ K<sup>+</sup> -ATPase inhibitor | No | - |
| -93.97 | periplocymarin | Na<sup>+</sup>/ K<sup>+</sup> -ATPase inhibitor | No | - |
| -93.34 | proscillaridin | Na<sup>+</sup>/ K<sup>+</sup> -ATPase inhibitor | No | - |
| -93.06 | thapsigargin | Ca<sup>2+</sup> -ATPase inhibitor | No | - |
| -92.92 | cinobufagin | Na<sup>+</sup>/ K<sup>+</sup> -ATPase inhibitor | Yes (Phase IV) | Liver cancer (NCT03843229) and gastrointestinal neoplasm (NCT02860429) |
| -92.46 | digitoxin | Na<sup>+</sup>/ K<sup>+</sup> -ATPase inhibitor | Yes (Phase II) | Cystic fibrosis (NCT00782288) and sarcoma (NCT00017446) |
| -92.35 | digoxin | Na<sup>+</sup>/ K<sup>+</sup> -ATPase inhibitor | Approved | Heart failure and atrial fibrillation |
| -92.33 | ouabain | Na<sup>+</sup>/ K<sup>+</sup> -ATPase inhibitor | No | - |
| -92.21 | bufalin | Na<sup>+</sup>/ K<sup>+</sup> -ATPase inhibitor | Yes (Phase II) | Pancreatic cancer (NCT00837239) |
| -90.84 | BNTX | Opioid receptor antagonist | No | - |

\*Connectivity scores $\leq -90$ provides compounds with opposing gene expression profiles.
FDA: Federal Drug Administration (USA); NCT numbers were obtained from ClinicalTrials.gov.

**Table 3.** Drug targets of TWAS-identified genes as identified by Open Targets Database.

| **Drug** | **Target protein** | **Indication** | **Status** |
| --- | --- | --- | --- |
| Letrozole | CYP19A1 | Breast cancer | Approved |
| Anastrozole | CYP19A1 | Breast cancer | Approved |
| Exemestane | CYP19A1 | Breast cancer | Approved |
| Aminoglutethimide | CYP19A1 | Breast cancer | Approved |
| Aminoglutethimide | CYP19A1 | Cushing syndrome | Approved |

## Discussion

We prioritised seven candidate susceptibility genes for endometrial cancer using a multi-tissue TWAS approach followed by colocalisation analysis. These genes included *AC145343*.*2*, located at a potentially novel endometrial cancer risk region which may be revealed by future better-powered GWAS. A secondary analysis revealed a further potential candidate susceptibility gene through association of imputed expression of *RP11-358N4*.*6* from testis with endometrioid endometrial cancer risk; however, this finding may not be robust because only a single, male-specific, tissue could genetically predict expression of this gene.

Two of the candidate susceptibility genes encode lncRNAs. Of these only *AC145343*.*2* appears to have been previously studied and is predicted to regulate apoptotic genes such as *CASP8* and *TNFRSF1A* (Liang et al., 2020). Consistent with an association between increased imputed expression of *AC145343*.*2* and increased endometrial cancer risk, we observed that *AC145343*.*2* is upregulated in The Cancer Genome Atlas endometrial tumours compared to normal endometrial or uterine tissue (http://gepia2.cancer-pku.cn/#analysis). Recently, we have identified the candidate susceptibility genes *BHLHE41, RAB11FIP4, SKAP1* and *SRCIN1* as potential regulatory targets of endometrial cancer GWAS risk variation through enhancer-promoter chromatin looping studies (O’Mara, Spurdle, Glubb, & Endometrial Cancer Association, 2019). Additionally, the TWAS-identified genes *EIF2AK4* and *SNX11*, which were not prioritised by colocalisation, had also been established as potential regulatory targets of endometrial cancer GWAS risk variation (O’Mara, Spurdle, Glubb, & Endometrial Cancer Association, 2019). The current study thus provides a crucial link between candidate GWAS target genes and endometrial cancer risk through their imputed expression.

Among these candidate susceptibility genes, *SRCIN1* (Src kinase signalling inhibitor 1) is a compelling biological candidate. Decreased *SRCIN1* expression was associated with increased endometrial cancer risk in our TWAS analysis. This finding is consistent with the tumour suppressor function of the protein encoded by *SRCIN1* which suppresses Src kinase activity (Cao et al., 2014; Wang et al., 2016; Xu et al., 2015; F. Yang et al., 2017) and the observation that Src kinase expression is increased in endometrial cancer (reviewed in (Grundker, Gunthert, & Emons, 2008)). Consistent with these observations, dasatinib, a Src kinase inhibitor, is being studied in clinical trials of recurrent endometrial cancer (ClinicalTrials.gov identifiers: NCT02059265 and NCT01440998).

In terms of the relationships between the candidate susceptibility genes and endometrial cancer, the best characterised gene is *CYP19A1*. We have previously linked *CYP19A1* to endometrial cancer susceptibility through genetic association with circulating estrogen (Thompson et al., 2016). *CYP19A1* encodes the aromatase enzyme, responsible for a rate-limiting step in the synthesis of estrogen and unopposed estrogen exposure is one of the most well established risk factors for endometrial cancer (Ignatov & Ortmann, 2020). Consistent with its function, increased *CYP19A1* expression was associated with increased endometrial cancer risk in our study. Furthermore, drug repurposing analysis linked *CYP19A1* to aromatase inhibitors (letrozole, anastrozole, exemestane and aminoglutethimide) which are used to treat breast cancer. Notably, letrozole has shown efficacy in treating endometrial hyperplasia (Moradan, Nikkhah, & Mirmohammadkhanai, 2017; Tabatabaie et al., 2013), a precursor to endometrial cancer; however, there is less evidence for the use of aromatase inhibitors to treat advanced endometrial cancer (Lindemann et al., 2014; Ma et al., 2004; Rose et al., 2000).

Phenome-wide lookup of candidate endometrial cancer susceptibility genes revealed associations with traits belonging to bone, hormone and diabetes phenotypic categories, which contain traits already related to endometrial cancer risk (O’Mara, Glubb, et al., 2019). Another phenotypic category contained cardiovascular traits (e.g. ischaemic heart disease and systolic blood pressure) which are associated with genetically established endometrial cancer risk factors such as body mass index, LDL and HDL cholesterol (Kho et al., 2020; O’Mara, Glubb, et al., 2019). These findings suggest that the identified traits belonging to the remaining phenotypic categories (liver function and haematopoiesis) may provide novel insights into endometrial cancer aetiology. GWAS data for these traits could be used to assess their effects on endometrial cancer risk using similar Mendelian randomisation approaches to our studies of body mass index and blood lipids (Kho et al., 2020; O’Mara, Glubb, et al., 2019).

We found 14 compounds that may counteract gene expression changes associated with endometrial cancer susceptibility. As drug targets supported by genetic evidence of disease association are more likely to receive clinical approval (King, Davis, & Degner, 2019; Nelson et al., 2015), there is a strong rationale for future pre-clinical endometrial cancer sensitivity studies of candidate compounds identified in this analysis. Six of the 14 compounds have either been clinically studied or approved for disease treatment and thus may have potential for drug repurposing. It is noteworthy that the two drugs with the strongest opposing gene expression profiles were the tubulin inhibitors nocodazole and vincristine, as the tubulin inhibitor paclitaxel is currently used to treat endometrial cancer patients with advanced disease (Burke et al., 2014). Although nocodazole does not appear to have been studied in humans, vincristine has been shown to have a limited clinical benefit on endometrial cancer but is associated with significant toxicity (Broun et al., 1993). Nine of the compounds were Na^+/^ K^+^-ATPase inhibitors, of which one, digoxin, has been approved for treatment of cardiac disease. Na^+^/K^+^-ATPase inhibitors have demonstrated anti-cancer activity in experimental models, including bufalin which was identified in our analysis and has shown effects specific to ovarian and endometrial cancer cell lines (Takai, Ueda, Nishida, Nasu, & Narahara, 2008). Although clinical studies have shown limited efficacy of Na^+^/K^+^-ATPase in the treatment of cancer, effects on endometrial cancer do not appear to have been studied yet (Durlacher et al., 2015). Cinacalcet, a calcium channel activator, approved for treatment of hypercalcemia and secondary hyperparathyroidism, has demonstrated anti-cancer activity in animal models (Rodríguez-Hernández et al., 2016) but has not yet been clinically studied as a cancer treatment. The Ca^2+^-ATPase inhibitor thapsigargin has demonstrated strong anti-proliferative effects in endometrial cancer cell lines in the Genomics of Drug Sensitivity in Cancer database (W. Yang et al., 2013). However, thapsigargin is not selective for cancer cells and thus a tumour-targeted derivative (mipsagargin) has been developed (Cui, Merritt, Fu, & Pan, 2017). Mipsagargin has been clinically studied for treatment of several tumour types (not including endometrial cancer) and shown modest activity (Mahalingam et al., 2019; Mahalingam et al., 2016).

We acknowledge some limitations to this study. As our TWAS analysis was performed using eQTL data from multiple tissues to maximise our power to detect candidate susceptibility genes, genes with tissue-specific expression may have been discounted. However, we have analysed the relatively small GTEx uterine eQTL sample set, arguably the most relevant tissue, and failed to identify genes that passed the TWAS multiple testing correction threshold. Furthermore, although we have applied colocalization analysis to identify concordant GWAS and eQTL signals and reduce false positive findings, the extent to which colocalization results represent genuine causal relationships remains unclear. Indeed, while we had previously established five of the seven genes with concordant signals as candidate endometrial cancer susceptibility genes or GWAS targets, two genes with discordant GWAS and eQTL signals also had evidence of targeting by endometrial cancer risk GWAS variation. Further functional studies will be necessary to determine the biological effects of the candidate susceptibility genes identified in this study on experimental phenotypes related to endometrial cancer risk.

In summary, our study has highlighted the value of using a multi-tissue transcriptome-wide association study and colocalisation analysis to prioritise biologically relevant candidate endometrial cancer susceptibility genes. As well as generating insights into biological mechanisms underlying endometrial cancer risk, we have identified compounds with potential for endometrial cancer repurposing and provided avenues for future studies of endometrial cancer aetiology.

## Methods

### Endometrial cancer risk GWAS data

We used summary data from the largest endometrial cancer GWAS meta-analysis to date, comprising 12,906 cases and 108,979 controls of European ancestry (O’Mara et al., 2018). Endometrial cancer subtype analyses were additionally performed using GWAS summary data restricting cases to endometrioid endometrial cancer patients (8,758 cases and 46,126 controls) (https://www.ebi.ac.uk/gwas/publications/30093612). For analysis, only variants located outside major histocompatibility complex region (chr6:26Mb–34Mb), were used.

### eQTL data

We used cis-eQTL summary statistics from 48 tissues that are available in GTEx Project (version 7). Cis-eQTL summary statistics for 48 tissues and information about respective sample sizes are available from the GTEx website (https://gtexportal.org/home/).

### Multi-tissue TWAS analysis

Multi-tissue TWAS analysis was performed in two steps. Firstly, we used S-PrediXcan (Barbeira et al., 2019) to incorporate endometrial cancer GWAS and eQTL data from each of the 48 GTEx tissues (version 7) and identify genes associated with endometrial cancer risk. Pre-specified weights based on covariance of genetic variants were used in S-PrediXcan analysis and obtained from PredictDB data repository (http://predictdb.org/). Secondly, S-PrediXcan data from all tissues were jointly analysed using multivariate regression in S-MultiXcan (Barbeira et al., 2019). To identify statistically significant TWAS genes from S-MultiXcan, a Bonferroni correction was considered. However, this approach would assume independent gene expression, whereas genes tend to be co-expressed (Wainberg et al., 2019). Therefore, we used a Benjamini-Hochberg false discovery rate (FDR) of less than 0.05 to correct for multiple comparisons.

### Colocalization analysis

Colocalization analysis was performed for each S-MultiXcan identified gene to assess the co-occurrence of endometrial cancer risk and eQTL signals from the best performing tissue, i.e. the most-significant tissue identified by S-PrediXcan. This approach enabled us to tease apart pleiotropic associations (where the same genetic variant is associated with gene expression and disease risk) from linkage (where correlated genetic variants are associated with gene expression and disease risk independently). Colocalization analysis was performed using the COLOC package with default parameters in R (Giambartolomei et al., 2014) and the input was identical to the TWAS analysis but restricted to variants within 1 Mb of the S-MultiXcan identified genes. COLOC provided posterior probabilities for the GWAS and eQTL signals to be explained by the same genetic variant. We considered a posterior probability greater than 0.75 for pleiotropic association as evidence of colocalization within a locus.

### Phenome-wide association analysis

We assessed pleiotropic associations of the TWAS-identified genes that showed evidence of colocalization by performing a phenome-wide lookup in the CTG-VIEW database (Cuéllar-Partida et al., 2019). The phenome-wide lookup was performed using summary-data-based Mendelian randomization (SMR) (Zhu et al., 2016) results from ∼1,600 phenotypes that are available from this platform. SMR analyses were performed using eQTL data from relevant or well-powered tissues (i.e. adipose subcutaneous, adipose visceral momentum, uterus and whole blood) and included a heterogeneity test (HEIDI) to identify candidate genes that are affected by the same GWAS risk variant, comparable to our colocalization analysis. Genes passing a Bonferroni correction (P_SMR_<7.2×10^−3^) and no evidence of heterogeneity (P_HEIDI_ > 0.05) were defined as having pleiotropic associations with candidate susceptibility genes.

### Computational drug repurposing analysis

We used the Connectivity Map platform (Subramanian et al., 2017) to identify compounds with gene expression profiles opposing the endometrial cancer risk TWAS data. As Connectivity Map analysis requires at least ten upregulated and ten downregulated coding genes as input, we used the identified TWAS genes from S-MultiXcan at FDR<0.15 (**Supplementary Table 3**). Connectivity scores were generated based on a modified Kolmogorov-Smirnov score, which summarises the relationship of the endometrial cancer TWAS gene expression profile with the drug-induced gene expression profile across cancer cell types in the Connectivity Map database. A connectivity score ≤ −90 suggest that the expression of the query genes opposes the drug-induced gene expression profile. We also used the Open Targets platform (Carvalho-Silva et al., 2019) to determine if candidate endometrial cancer susceptibility genes encode known targets for drugs that have been clinically studied.

## Data Availability

Summary-level GWAS meta-analysis results for endometrial cancer that support the findings of this study are available at the NHGRI-EBI GWAS Catalog (https://www.ebi.ac.uk/gwas/downloads/summary-statistics). Cis-eQTL summary statistics for 48 tissues are available at the GTEx website (https://gtexportal.org/home/). Other data generated and/or analyzed during this study are included in this article and its supplementary information files or are available on reasonable request.

## Acknowledgements

We thank the many women who participated in the Endometrial Cancer Association Consortium, and the numerous institutions and their staff who supported recruitment. A full list of consortium members and acknowledgements can be found in the Supplementary Note.

