## Supplementary Materials (figures and tables) for "Multi-tissue transcriptome-wide association study identifies genetic mechanisms underlying endometrial cancer susceptibility"

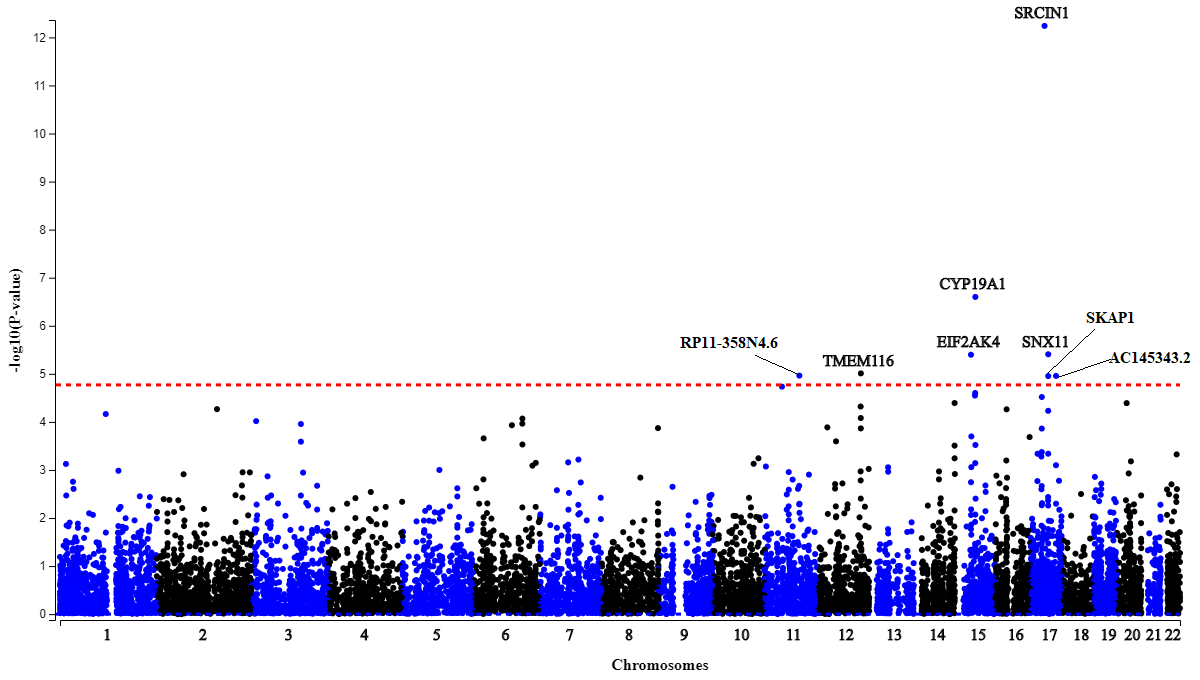

**Supplementary Figure 1. Manhattan plot of TWAS analysis of endometrioid endometrial cancer risk. Genes are plotted according to chromosome and position against -log_10_(P value) of TWAS association.** Red dotted lines represent Benjamini-Hochberg FDR<0.05 significance threshold, which is equivalent to P<1.7 ×10^-5^.

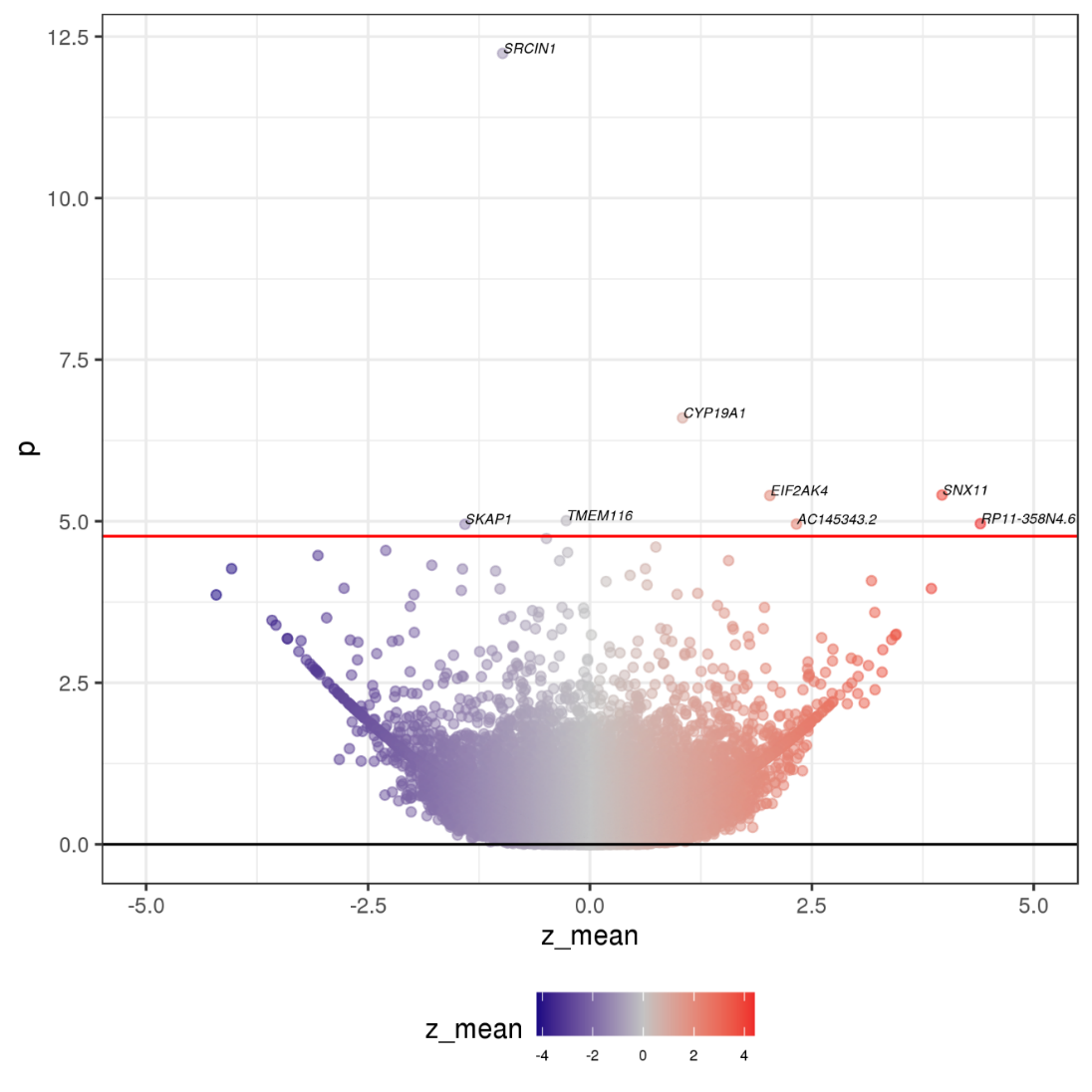

**Supplementary Figure 2. Volcano plot from TWAS analysis of endometrioid endometrial cancer risk.** Each dot represents the effect of the average predicted expression of each gene on endometrial cancer risk. Red line represents the Benjamini-Hochberg FDR<0.05 significance threshold.

**Supplementary Table 1. List of genes associated with endometrioid endometrial cancer risk (FDR < 0.05)**

| Region | Gene | Best performing tissue | N_tissue_ | P | FDR | Z_min_ | Z_max_ | Z_mean_ | Z_SD_ | PP_COLOC_ |
| --- | --- | --- | --- | --- | --- | --- | --- | --- | --- | --- |
| 17q12 | ***SRCIN1^#^*** | Esophagus Gastroesophageal Junction | 6 | 5.76E-13 | 1.47E-08 | -7.62 | 2.03 | -0.98 | 3.54 | 0.848 |
| 15q21.2 | ***CYP19A1^#^*** | Skin Sun Exposed Lower leg | 8 | 2.51E-07 | 3.20E-03 | -2.71 | 5.57 | 1.04 | 2.67 | 0.987 |
| 17q21.32 | ***SNX11*** | Artery Tibial | 26 | 3.93E-06 | 2.55E-02 | 0.84 | 5.46 | 3.97 | 1.14 | 0.808 |
| 15q15.1 | *EIF2AK4* | Colon Transverse | 31 | 4.00E-06 | 2.55E-02 | -2.09 | 5.01 | 2.03 | 1.97 | 0.115 |
| 12q24.13 | *TMEM116* | Lung | 24 | 9.80E-06 | 3.55E-02 | -4.01 | 1.08 | -0.27 | 1.23 | 0.003 |
| 11q14.3 | ***RP11-358N4.6*** | Testis | 1 | 1.09E-05 | 3.55E-02 | 4.40 | 4.40 | 4.40 | NA | 0.771 |
| 17q24.2 | ***AC145343.2*** | Adipose Visceral Omentum | 11 | 1.11E-05 | 0.03548 | -0.58 | 4.44 | 2.33 | 1.78 | 0.845 |
| 17q21.32 | ***SKAP1*** | Liver | 8 | 1.11E-05 | 0.03548 | -5.49 | 0.74 | -1.41 | 1.99 | 0.930 |

N_tissue_: number of tissues available for this gene; FDR: false discovery rate; Z_min_: minimum Z score from single tissue S-PrediXcan result; Z_max_: maximum Z score from single tissue S-PrediXcan result; Z_mean_: mean Z score from single tissue S-PrediXcan result; Z_SD_: standard deviation of Z score from single tissue S-PrediXcan result. PP_COLOC_: posterior probability that endometrial cancer risk and eQTL variants from best performing tissue colocalize. *^#^*TWAS association passed Bonferroni correction threshold (P<2×10^-6^). **Genes in bold** showed evidence of colocalization (PP_COLOC_>0.75). NA: not available.

**Supplementary Table 2. Phenotypes associated with candidate endometrial cancer susceptibility genes**

| Categories | Phenotypes | Genes | P value |
| --- | --- | --- | --- |
| Bone | Ankle spacing width | *AC145343.2* | 5.14E-05 |
|  | Bone fracture | *AC145343.2* | 2.80E-04 |
|  | Bone mineral density | *AC145343.2* | 2.45E-03 |
|  | Gonarthrosis | *RAB11FIP4* | 9.60E-04 |
|  | Heel bone mineral density | *AC145343.2* | 1.52E-03 |
|  | Heel bone mineral density | *CYP19A1* | 2.73E-07 |
|  | Heel bone mineral density | *RAB11FIP4* | 2.99E-04 |
| Cardiovascular | Ischaemic heart disease | *RAB11FIP4* | 5.24E-03 |
|  | Self-reported heart arrhythmia | *CYP19A1* | 3.52E-03 |
|  | Systolic blood pressure | *AC145343.2* | 4.95E-03 |
|  | Treatment/medication code: candesartan cilexetil | *CYP19A1* | 3.72E-04 |
| Haematopoiesis | Mean corpuscular volume | *AC145343.2* | 5.46E-03 |
|  | Mean platelet volume | *RAB11FIP4* | 5.15E-03 |
|  | Myeloid white cell count | *CYP19A1* | 2.99E-03 |
|  | Neutrophil count | *CYP19A1* | 5.11E-03 |
|  | Neutrophil percentage | *CYP19A1* | 2.07E-04 |
|  | Platelet count | *CYP19A1* | 5.46E-03 |
|  | Platelet crit | *CYP19A1* | 1.50E-03 |
|  | Platelet distribution width | *AC145343.2* | 2.48E-03 |
|  | Sum neutrophil eosinophil counts | *CYP19A1* | 2.99E-03 |
|  | White blood cell count | *CYP19A1* | 3.97E-03 |
| Diabetes | Glucose | *RAB11FIP4* | 3.55E-03 |
|  | Glycated haemoglobin | *AC145343.2* | 8.10E-04 |
|  | Glycated haemoglobin | *CYP19A1* | 3.89E-03 |
|  | IGF-1 | *CYP19A1* | 1.81E-05 |
|  | Treatment/medication code: Gliclazide | *AC145343.2* | 2.16E-03 |
| Sex hormones | Age at last live birth | *AC145343.2* | 9.67E-04 |
|  | Age at menopause | *AC145343.2* | 1.61E-04 |
|  | Testosterone | *CYP19A1* | 1.92E-05 |
| Liver | Albumin | *CYP19A1* | 4.09E-03 |
|  | Alkaline phosphatase | *CYP19A1* | 4.20E-05 |
|  | Direct bilirubin | *RAB11FIP4* | 2.94E-03 |
|  | Direct bilirubin | *AC145343.2* | 4.94E-03 |
|  | Disorders of gallbladder, biliary tract and pancreas | *RAB11FIP4* | 4.79E-03 |
|  | Gamma glutamyltransferase | *RAB11FIP4* | 4.41E-03 |

**Supplementary Table 3. Genes associated with endometrial cancer risk (FDR<0.15)**

| Gene | Best performing tissue | N_tissue_ | P | FDR | Z_min_ | Z_max_ | Z_mean_ | Z_SD_ |
| --- | --- | --- | --- | --- | --- | --- | --- | --- |
| *SRCIN1* | Esophagus Gastroesophageal Junction | 6 | 9.49E-17 | 2.42E-12 | -8.96 | 2.00 | -1.59 | 4.10 |
| *CYP19A1* | Skin Sun Exposed Lower leg | 8 | 2.93E-08 | 3.73E-04 | -2.31 | 5.76 | 1.34 | 2.89 |
| *EIF2AK4* | Colon Transverse | 31 | 5.19E-07 | 4.41E-03 | -2.56 | 5.44 | 2.41 | 1.97 |
| *IQSEC1* | Esophagus Mucosa | 11 | 7.81E-07 | 4.97E-03 | -1.91 | 4.76 | 0.73 | 1.92 |
| *BHLHE41* | Thyroid | 8 | 1.23E-06 | 6.25E-03 | -1.09 | 5.82 | 2.05 | 2.28 |
| *RP11-521C20.2* | Heart Left Ventricle | 6 | 2.16E-06 | 9.18E-03 | -1.30 | 5.39 | 1.93 | 2.74 |
| *HINT3* | Esophagus Muscularis | 3 | 4.20E-06 | 0.01 | -4.21 | 3.36 | 0.66 | 4.23 |
| *GLDN* | Adipose Subcutaneous | 21 | 4.38E-06 | 0.01 | -2.40 | 3.75 | -0.68 | 1.43 |
| *TMEM116* | Lung | 24 | 5.68E-06 | 0.02 | -4.19 | 1.18 | -0.37 | 1.27 |
| *SPPL2A* | Stomach | 30 | 6.27E-06 | 0.02 | -3.05 | 2.75 | 0.95 | 1.11 |
| *AC145343.2* | Esophagus Muscularis | 11 | 7.85E-06 | 0.02 | -0.61 | 4.55 | 2.47 | 1.91 |
| *TRMT11* | Stomach | 5 | 1.10E-05 | 0.02 | -3.24 | 5.04 | -1.24 | 3.53 |
| *RHOV* | Brain Cerebellum | 6 | 1.59E-05 | 0.03 | -1.91 | 3.07 | 1.54 | 2.25 |
| *RP11-1407O15.2* | Brain Anterior cingulate cortex BA24 | 10 | 1.95E-05 | 0.03 | -5.84 | 1.95 | -0.29 | 2.06 |
| *SNX11* | Adipose Subcutaneous | 26 | 2.01E-05 | 0.03 | 1.27 | 5.59 | 4.11 | 0.98 |
| *RAB11FIP4* | Esophagus Mucosa | 12 | 2.05E-05 | 0.03 | -2.72 | 3.51 | -0.26 | 2.40 |
| *LINGO1* | Esophagus Muscularis | 16 | 2.27E-05 | 0.03 | -1.61 | 4.78 | 1.38 | 1.62 |
| *RP1-46F2.2* | Whole Blood | 5 | 3.30E-05 | 0.05 | -3.90 | -2.25 | -3.01 | 0.72 |
| *SKAP1* | Liver | 8 | 3.79E-05 | 0.05 | -5.68 | 0.98 | -1.46 | 1.91 |
| *RP11-57A19.4* | Lung | 1 | 3.84E-05 | 0.05 | -4.12 | -4.12 | -4.12 | NA |
| *SEC61A1* | Cells Transformed fibroblasts | 5 | 3.85E-05 | 0.05 | -4.09 | 1.40 | -1.16 | 2.17 |
| *SMURF2P1* | Cells Transformed fibroblasts | 4 | 3.93E-05 | 0.05 | -2.58 | 2.58 | 0.91 | 2.35 |
| *AC138744.2* | Adrenal Gland | 1 | 5.28E-05 | 0.06 | -4.04 | -4.04 | -4.04 | NA |
| *GABPB1* | Artery Tibial | 3 | 5.29E-05 | 0.06 | -4.22 | 0.20 | -2.21 | 2.24 |
| *GS1-259H13.2* | Small Intestine Terminal Ileum | 28 | 6.10E-05 | 0.06 | -0.94 | 2.81 | 1.24 | 1.22 |
| *EVI2A* | Esophagus Mucosa | 7 | 6.69E-05 | 0.07 | -4.90 | -3.34 | -4.33 | 0.59 |
| *MVD* | Lung | 5 | 7.67E-05 | 0.07 | -3.41 | 0.43 | -1.99 | 1.50 |
| *EEFSEC* | Lung | 18 | 7.68E-05 | 0.07 | 0.61 | 4.87 | 3.59 | 1.37 |
| *HEY2* | Ovary | 13 | 8.91E-05 | 0.08 | -4.92 | -0.87 | -2.97 | 1.03 |
| *ALDH2* | Esophagus Muscularis | 17 | 1.03E-04 | 0.09 | -3.87 | 1.19 | -2.30 | 1.19 |
| *RP11-1348G14.4* | Colon Transverse | 40 | 1.21E-04 | 0.10 | -3.80 | -0.72 | -2.78 | 0.87 |
| *TUFM* | Artery Aorta | 36 | 1.29E-04 | 0.10 | -0.22 | 3.60 | 2.63 | 0.96 |
| *COL6A2* | Whole Blood | 9 | 1.30E-04 | 0.10 | -2.87 | 2.99 | 0.19 | 1.71 |
| *UQCC1* | Artery Tibial | 21 | 1.32E-04 | 0.10 | -4.68 | 0.10 | -3.50 | 1.00 |
| *ESR1* | Testis | 3 | 1.39E-04 | 0.10 | 0.36 | 4.35 | 1.86 | 2.17 |
| *SPN* | Stomach | 2 | 1.43E-04 | 0.10 | -4.00 | 0.02 | -1.99 | 2.85 |
| *HIPK4* | Colon Transverse | 11 | 1.51E-04 | 0.10 | -3.78 | -0.58 | -2.23 | 1.11 |
| *DCAF13P3* | Whole Blood | 5 | 1.62E-04 | 0.11 | 0.00 | 3.85 | 1.40 | 1.70 |
| *ENTPD6* | Testis | 25 | 1.72E-04 | 0.11 | -3.84 | 1.14 | -1.11 | 1.26 |
| *NFE2L1* | Esophagus Mucosa | 7 | 1.73E-04 | 0.11 | -4.77 | 3.19 | -1.21 | 2.48 |
| *BCL7B* | Cells Transformed fibroblasts | 11 | 1.89E-04 | 0.12 | -3.22 | 0.38 | -2.27 | 1.21 |
| *RP11-81M19.3* | Testis | 2 | 1.93E-04 | 0.12 | -3.84 | -3.77 | -3.81 | 0.05 |
| *SRP14-AS1* | Brain Spinal cord cervical c-1 | 38 | 2.04E-04 | 0.12 | -3.81 | 1.06 | -0.26 | 1.14 |
| *LY6E* | Testis | 9 | 2.10E-04 | 0.12 | -2.73 | 3.39 | 1.36 | 2.05 |
| *POU3F2* | Brain Cerebellar Hemisphere | 2 | 2.15E-04 | 0.12 | -4.04 | 0.94 | -1.55 | 3.52 |
| *ADSSL1* | Testis | 15 | 2.47E-04 | 0.13 | -4.06 | 4.14 | -2.74 | 2.22 |
| *RUVBL1* | Cells Transformed fibroblasts | 6 | 2.52E-04 | 0.13 | -3.21 | 2.36 | 0.13 | 2.08 |
| *BRAP* | Skin Not Sun Exposed Suprapubic | 3 | 2.55E-04 | 0.13 | 1.25 | 4.31 | 3.02 | 1.59 |
| *PRDM5* | Cells Transformed fibroblasts | 25 | 2.59E-04 | 0.13 | -2.92 | 0.67 | -1.59 | 1.07 |

N_tissue_: number of tissues available for this gene; FDR: false discovery rate; Z_min_: minimum Z score from single tissue S-PrediXcan result; Z_max_: maximum Z score from single tissue S-PrediXcan result; Z_mean_: mean Z score from single tissue S-PrediXcan result; Z_SD_: standard deviation of Z score from single tissue S-PrediXcan result.
