## Supplementary Note for "Multi-tissue transcriptome-wide association study identifies genetic mechanisms underlying endometrial cancer susceptibility"

**Endometrial Cancer Association Consortium Collaborators**

Frederic Amant^1^, Daniela Annibali^1^, Katie Ashton^2-4^, John Attia^2, 5^, Paul L. Auer^6, 7^, Matthias W. Beckmann^8^, Amanda Black^9^, Louise Brinton^9^, Daniel D. Buchanan^10-13^, Stephen J. Chanock^14^, Chu Chen^15^, Maxine M. Chen^16^, Timothy H.T. Cheng^17^, Linda S. Cook^18, 19^, Marta Crous-Bous^16, 20^, Kamila Czene^21^, Immaculata De Vivo^16, 20^, Joe Dennis^22^, Thilo Dörk^23^, Sean C. Dowdy^24^, Alison M. Dunning^25^, Matthias Dürst^26^, Douglas F. Easton^22, 25^, Arif B. Ekici^27^, Peter A. Fasching^8, 28^, Brooke L. Fridley^29^, Christine M. Friedenreich^19^, Montserrat García-Closas^14^, Mia M. Gaudet^30^, Graham G. Giles^11, 31, 32^, Dylan M. Glubb^33^, Ellen L. Goode^34^, Christopher A. Haiman^35^, Per Hall^21, 36^, Susan E. Hankinson^20, 37^, Catherine S. Healey^25^, Alexander Hein^8^, Peter Hillemanns^23^, Shirley Hodgson^38^, Erling Hoivik^39, 40^, Elizabeth G. Holliday^2, 5^, David J. Hunter^16, 41^, Angela Jones^17^, Peter Kraft^16, 42^, Camilla Krakstad^39, 40^, Diether Lambrechts^43, 44^, Loic Le Marchand^45^, Xiaolin Liang^46^, Annika Lindblom^47, 48^, Jolanta Lissowska^49^, Jirong Long^50^, Lingeng Lu^51^, Anthony M. Magliocco^52^, Lynn Martin^53^, Mark McEvoy^5^, Roger L. Milne^11, 31, 32^, Miriam Mints^54^, Rami Nassir^55^, Tracy A. O'Mara^33^, Irene Orlow^46^, Geoffrey Otton^56^, Claire Palles^17^, Paul D.P. Pharoah^22, 25^, Loreall Pooler^35^, Tony Proietto^56^, Timothy R. Rebbeck^57, 58^, Stefan P. Renner^59^, Harvey A. Risch^51^, Matthias Rübner^59^, Ingo Runnebaum^26^, Carlotta Sacerdote^60, 61^, Gloria E. Sarto^62^, Fredrick Schumacher^63^, Rodney J. Scott^2, 4, 64^, V. Wendy Setiawan^35^, Mitul Shah^25^, Xin Sheng^35^, Xiao-Ou Shu^50^, Melissa C. Southey^10, 31, 32^, Amanda B. Spurdle^33^, Emma Tham^47, 65^, Deborah J. Thompson^22^, Ian Tomlinson^17, 53^, Jone Trovik^39, 40^, Constance Turman^16^, David Van Den Berg^35^, Zhaoming Wang^9^, Penelope M. Webb^66^, Nicolas Wentzensen^9^, Stacey J. Winham^67^, Lucy Xia^35^, Yong-Bing Xiang^68^, Hannah P. Yang^9^, Herbert Yu^45^, Wei Zheng^50^

^1^ Department of Obstetrics and Gynecology, Division of Gynecologic Oncology, University Hospitals KU Leuven, University of Leuven, Leuven, Belgium. ^2^ Hunter Medical Research Institute, John Hunter Hospital, Newcastle, New South Wales, Australia. ^3^ Centre for Information Based Medicine, University of Newcastle, Callaghan, New South Wales, Australia.
^4^ Discipline of Medical Genetics, School of Biomedical Sciences and Pharmacy, Faculty of Health, University of Newcastle, Callaghan, New South Wales, Australia. ^5^ Centre for Clinical Epidemiology and Biostatistics, School of Medicine and Public Health, University of Newcastle, Callaghan, New South Wales, Australia. ^6^ Cancer Prevention Program, Fred Hutchinson Cancer Research Center, Seattle, WA, USA. ^7^ Zilber School of Public Health, University of Wisconsin-Milwaukee, Milwaukee, WI, USA. ^8^ Department of Gynecology and Obstetrics, Comprehensive Cancer Center ER-EMN, University Hospital Erlangen, Friedrich-Alexander-University Erlangen-Nuremberg, Erlangen, Germany. ^9^ Division of Cancer Epidemiology and Genetics, National Cancer Institute, Bethesda, MD, USA. ^10^ Department of Clinical Pathology, The University of Melbourne, Melbourne, Victoria, Australia. ^11^ Centre for Epidemiology and Biostatistics, Melbourne School of Population and Global Health, The University of Melbourne, Melbourne, Victoria, Australia. ^12^ Genomic Medicine and Family Cancer Clinic, Royal Melbourne Hospital, Parkville, Victoria, Australia. ^13^ University of Melbourne Centre for Cancer Research, Victorian Comprehensive Cancer Centre, Parkville, Victoria, Australia. ^14^ Division of Cancer Epidemiology and Genetics, National Cancer Institute, National Institutes of Health, Department of Health and Human Services, Bethesda, MD, USA. ^15^ Epidemiology Program, Fred Hutchinson Cancer Research Center, Seattle, WA, USA. ^16^ Department of Epidemiology, Harvard T.H. Chan School of Public Health, Boston, MA, USA. ^17^ Wellcome Trust Centre for Human Genetics and Oxford NIHR Biomedical Research Centre, University of Oxford, Oxford, UK. ^18^ University of New Mexico Health Sciences Center, University of New Mexico, Albuquerque, NM, USA. ^19^ Department of Cancer Epidemiology and Prevention Research, Alberta Health Services, Calgary, AB, Canada. ^20^ Channing Division of Network Medicine, Department of Medicine, Brigham and Women's Hospital and Harvard Medical School, Boston, MA, USA. ^21^ Department of Medical Epidemiology and Biostatistics, Karolinska Institutet, Stockholm, Sweden. ^22^ Centre for Cancer Genetic Epidemiology, Department of Public Health and Primary Care, University of Cambridge, Cambridge, UK. ^23^ Gynaecology Research Unit, Hannover Medical School, Hannover, Germany. ^24^ Department of Obstetrics and Gynecology, Division of Gynecologic Oncology, Mayo Clinic, Rochester, MN, USA. ^25^ Centre for Cancer Genetic Epidemiology, Department of Oncology, University of Cambridge, Cambridge, UK.
^26^ Department of Gynaecology, Jena University Hospital - Friedrich Schiller University, Jena, Germany. ^27^ Institute of Human Genetics, University Hospital Erlangen, Friedrich-Alexander University Erlangen-Nuremberg, Comprehensive Cancer Center Erlangen-EMN, Erlangen, Germany. ^28^ David Geffen School of Medicine, Department of Medicine Division of Hematology and Oncology, University of California at Los Angeles, Los Angeles, CA, USA.
^29^ Department of Biostatistics, Kansas University Medical Center, Kansas City, KS, USA.
^30^ Department of Population Science, American Cancer Society, Atlanta, GA, USA. ^31^ Cancer Epidemiology Division, Cancer Council Victoria, Melbourne, Victoria, Australia. ^32^ Precision Medicine, School of Clinical Sciences at Monash Health, Monash University, Clayton, Victoria, Australia. ^33^ Department of Genetics and Computational Biology, QIMR Berghofer Medical Research Institute, Brisbane, Queensland, Australia. ^34^ Department of Health Science Research, Division of Epidemiology, Mayo Clinic, Rochester, MN, USA. ^35^ Department of Preventive Medicine, Keck School of Medicine, University of Southern California, Los Angeles, CA, USA. ^36^ Department of Oncology, Södersjukhuset, Stockholm, Sweden. ^37^ Department of Biostatistics & Epidemiology, University of Massachusetts, Amherst, Amherst, MA, USA. ^38^ Department of Clinical Genetics, St George's, University of London, London, UK. ^39^ Centre for Cancer Biomarkers CCBIO, Department of Clinical Science, University of Bergen, Bergen, Norway.
^40^ Department of Obstetrics and Gynecology, Haukeland University Hospital, Bergen, Norway. ^41^ Nuffield Department of Population Health, University of Oxford, Oxford, UK. ^42^ Program in Genetic Epidemiology and Statistical Genetics, Harvard T.H. Chan School of Public Health, Boston, MA, USA. ^43^ VIB Center for Cancer Biology, Leuven, Belgium. ^44^ Laboratory for Translational Genetics, Department of Human Genetics, University of Leuven, Leuven, Belgium. ^45^ Epidemiology Program, University of Hawaii Cancer Center, Honolulu, HI, USA.
^46^ Department of Epidemiology and Biostatistics, Memorial Sloan-Kettering Cancer Center, New York, NY, USA. ^47^ Department of Molecular Medicine and Surgery, Karolinska Institutet, Stockholm, Sweden. ^48^ Department of Clinical Genetics, Karolinska University Hospital, Stockholm, Sweden. ^49^ Department of Cancer Epidemiology and Prevention, M. Sklodowska-Curie Cancer Center, Oncology Institute, Warsaw, Poland. ^50^ Division of Epidemiology, Department of Medicine, Vanderbilt Epidemiology Center, Vanderbilt-Ingram Cancer Center, Vanderbilt University School of Medicine, Nashville, TN, USA. ^51^ Chronic Disease Epidemiology, Yale School of Public Health, New Haven, CT, USA. ^52^ Department of Anatomic Pathology, Moffitt Cancer Center & Research Institute, Tampa, FL, USA. ^53^ Institute of Cancer and Genomic Sciences, University of Birmingham, Birmingham, UK. ^54^ Department of Women's and Children's Health, Karolinska Institutet, Stockholm, Sweden. ^55^ Department of Biochemistry and Molecular Medicine, University of California Davis, Davis, CA, USA. ^56^ School of Medicine and Public Health, University of Newcastle, Callaghan, New South Wales, Australia. ^57^ Harvard T.H. Chan School of Public Health, Boston, MA, USA. ^58^ Dana-Farber Cancer Institute, Boston, MA, USA. ^59^ Department of Gynaecology and Obstetrics, University Hospital Erlangen, Friedrich-Alexander University Erlangen-Nuremberg, Comprehensive Cancer Center Erlangen-EMN, Erlangen, Germany. ^60^ Center for Cancer Prevention (CPO-Peimonte), Turin, Italy. ^61^ Human Genetics Foundation (HuGeF), Turino, Italy. ^62^ Department of Obstetrics and Gynecology, School of Medicine and Public Health, University of Wisconsin, Madison, WI, USA. ^63^ Department of Population and Quantitative Health Sciences, Case Western Reserve University, Cleveland, OH, USA. ^64^ Division of Molecular Medicine, Pathology North, John Hunter Hospital, Newcastle, New South Wales, Australia. ^65^ Clinical Genetics, Karolinska Institutet, Stockholm, Sweden. ^66^ Population Health Department, QIMR Berghofer Medical Research Institute, Brisbane, Queensland, Australia. ^67^ Department of Health Sciences Research, Division of Biomedical Statistics and Informatics, Mayo Clinic, Rochester, MN, USA. ^68^ State Key Laboratory of Oncogene and Related Genes & Department of Epidemiology, Shanghai Cancer Institute, Renji Hospital, Shanghai Jiaotong University School of Medicine, Shanghai, China.

**Endometrial Cancer Association Consortium Acknowledgements and Funding**

We thank the participants in the endometrial cancer studies included in our study. The Endometrial Cancer Association Consortium genome-wide association analyses were supported by the National Health and Medical Research Council of Australia (APP552402, APP1031333, APP1109286, APP1111246 and APP1061779), the U.S. National Institutes of Health (R01-CA134958), European Research Council (EU FP7 Grant), Wellcome Trust Centre for Human Genetics (090532/Z/09Z) and Cancer Research UK. OncoArray genotyping of ECAC cases was performed with the generous assistance of the Ovarian Cancer Association Consortium (OCAC), which was funded through grants from the U.S. National Institutes of Health (CA1X01HG007491-01 (C.I. Amos), U19-CA148112 (T.A. Sellers), R01-CA149429 (C.M. Phelan) and R01-CA058598 (M.T. Goodman); Canadian Institutes of Health Research (MOP-86727 (L.E. Kelemen)) and the Ovarian Cancer Research Fund (A. Berchuck). We particularly thank the efforts of Cathy Phelan. OncoArray genotyping of the BCAC controls was funded by Genome Canada Grant GPH-129344, NIH Grant U19 CA148065, and Cancer UK Grant C1287/A16563. All studies and funders are listed in O’Mara et al (2018).
